# Metastatic Lymph Node Station Number Predicts Survival in Small Cell Lung Cancer

**DOI:** 10.1101/2020.09.06.20189191

**Authors:** Han Zhang, Cong Jiang, Kaiqi Jing, Jing Zhang, Yan Chen, Yuming Zhu, Gening Jiang, Peng Zhang

## Abstract

**Purpose:** As for pathologic N category, various regrouping strategies have been raised in non-small cell lung cancer (NSCLC) but little was done in small cell lung cancer(SCLC). On the basis of the suggestions discussed in NSCLC, we proposed a novel, metastatic lymph node station number (MNSN) - based pathologic N parameter and compared its efficacy in predicting survival with pN in SCLC.

**Methods:** We retrospectively analyzed the patients operated and pathologically diagnosed as SCLC in our hospital between 2009 and 2019. Kaplan–Meier method and Cox regression analysis were used to compare survival between groups defined by pN and MNSN.

**Results:** From 2009 to 2019, 566 patients received surgery for SCLC and 530 of them were eligible for subsequent analysis, with a median follow-up time of 21 months. The 5-year overall survival (OS) rates were 58.8%, 38.6%, 27.9% for pN0, pN1, pN2 stages and were 58.8%, 36.8%, 22.1%, 0% for MNSN0, 1-2, 3-5, 6-7 groups, respectively. Analyses of overall and recurrence-free survival (RFS) revealed that pN1 could not be distinguished from pN2 (OS, p = 0.099; RFS, p = 0.254), but the groups in MNSN were well separated from each other (OS, p< 0.001, p = 0.001, p = 0.063; RFS, p< 0.001, p = 0.026, p = 0.01, compared with the former group). When adjusted for sex, age, smoking, tumor purity and T stage, MNSN groups were independent hazard factors for OS and RFS.

**Conclusions:** Based on our cohort study, the MNSN-based N parameter might be a better indicator to predict survival than pN in SCLC and worth considering in the definition of N category in the future.

## Introduction

Lung cancer continues to be the leading cause of morbidity and mortality among all cancers^1^. Especially, SCLC which represents about 15% of lung cancers has worst prognosis^2,3^. SCLC is characterized by rapid growth rate, early dissemination to regional lymph nodes and distant sites. Although chemotherapy and radiotherapy are recommended for SCLC, the role of surgery still can’t be omitted^4^.

In clinical practice of cancers, accurate staging is just second to pathological classifications, which could guide the treatment schemes and predict survival. The staging project for SCLC has shifted from VALG staging system proposed in 1950s^5,6^ to TNM staging system in 2009^7,8^. However, since the release of seventh edition of the TNM staging system in 2009, the N parameter has not been changed in the last decade. The latest N parameter recommended by the eighth edition of TNM staging system^9^ is subdivided into N0, N1, N2 and N3 to represent the range of involved lymph node (LN) groups and is identical in SCLC^10^ and NSCLC^11^. In recent years, more and more studies have revealed the heterogeneities in N1 and N2 disease^12,13^. Accordingly, several parameters were put forward to regroup N1 or N2 stages in NSCLC, such as the number of positive LNs (nN)^14,15^, the ratio for the number of positive to sampled LNs (LNR)^16,17^, single or multiple-station metastasis^18,19^, presence of skip metastasis^11,20^, and their combination with pN^21,22^, et al.

Whereas, few studies were carried out to discuss the regrouping strategy of pathologic N stage in SCLC, probably due to the limited number of operated patients. Actually, the staging strategies for LN metastasis used in SCLC were originally proposed in NSCLC and then verified by SCLC database by International Association for the Study of Lung Cancer (IASLC)^8,10^. Although it was confirmed effective, it may not be ideal considering the difference in cytology, histology and invasiveness between SCLC and NSCLC. Based on the researches in NSCLC, we assumed that metastatic lymph node station number (MNSN) might be related to survival in SCLC.

The aim of this study was to verify the efficacy of MNSN in predicting survival in SCLC and compare it with the pN in eighth edition of TNM staging system.

## Materials and methods

### Study Design and Data Source

Patients who underwent pulmonary surgery and were pathologically diagnosed as SCLC in Shanghai Pulmonary Hospital, China from January 2009 to July 2019 were analyzed. Surgeries intended for curative resection with complete mediastinal LN dissection or systematic mediastinal LN sampling were performed. Postoperative chemotherapy with a platin-based regimen was routinely administered except for patients unendurable or refused. Postoperative radiotherapy was administered at the discretion of the radiation oncologists and the suggestion of the referring surgeons. Follow-up was executed by our staff regularly since the discharge from hospital.

Exclusion criteria were as follows: (1)Patients with a history of lung cancer; (2) Patients with metastasis; (3)Patients with R1 or R2 resection; (4)Patients without pathologic lymph nodes evaluation for any reasons; (5)Perioperative mortality; (6) Lose contact after being discharged from hospital.

The data of SCLC patients in The Surveillance, Epidemiology, and End Results (SEER) database was also used.

### Definitions

Overall survival (OS) is defined as the time between date of surgery and the date of death. Patients alive at last contact or lose connection during follow-up were censored at the last contact date. Recurrence free survival (RFS) is defined as the time between date of surgery and the date of cancer recurrence or last follow-up. The anatomical classification of LN stations is defined according to international LN map (IASLC map). The metastatic lymph node station number means the number of LN stations which has at least one node pathologically confirmed invasion of cancer cells.

### Statistical Analysis

Categorical variables were shown as frequencies and percentages, with γ^2^ tests being used for distribution analyses. Continuous variables were presented as mean ± standard deviation (SD) or median with ranges. Univariate and multivariate Cox proportional hazards model were employed to determine factors significantly associated with OS and RFS. Variables generated from univariate analysis (p < 0.1) or reported to be associated with survival were included in multivariate analysis (sex, age, smoking, tumor purity, pathologic T stage and MNSN group). Actuarial survival curves were plotted by the Kaplan-Meier method and the differences among groups were analyzed by log-rank test. All statistical analyses were two-sided tested and a probability value less than 0.05 was considered statistically significant. Analyses were performed using Statistical Package for Social Sciences software (SPSS, version 23, IBM Corporation, Armonk, NY).

## Results

### Baseline Characteristics

From 2009 to 2019, 566 patients were operated and pathologically confirmed SCLC. Among them, 547 patients had LN evaluation. 5 patients with M status, 1 patient with a history of lung cancer, 8 patients died perioperatively and 3 patients losing contact were excluded according to exclusion criteria. Eventually, 530 patients were included in this study (Fig. 1). The average age was 60.7 ± 9.0 years, with male making up 87.0%. Video-assisted thoracoscope surgery (VATS) were performed in 268 patients (50.6%), in which 100 patients (37.3%) received uniportal approach (Table 1). The median number of sampled lymph node station was 6 (range 1-11), and the median number of metastatic lymph node station was 1 (range 0-7). There were no patients in pathologic N3 stage.

**Figure 1.**
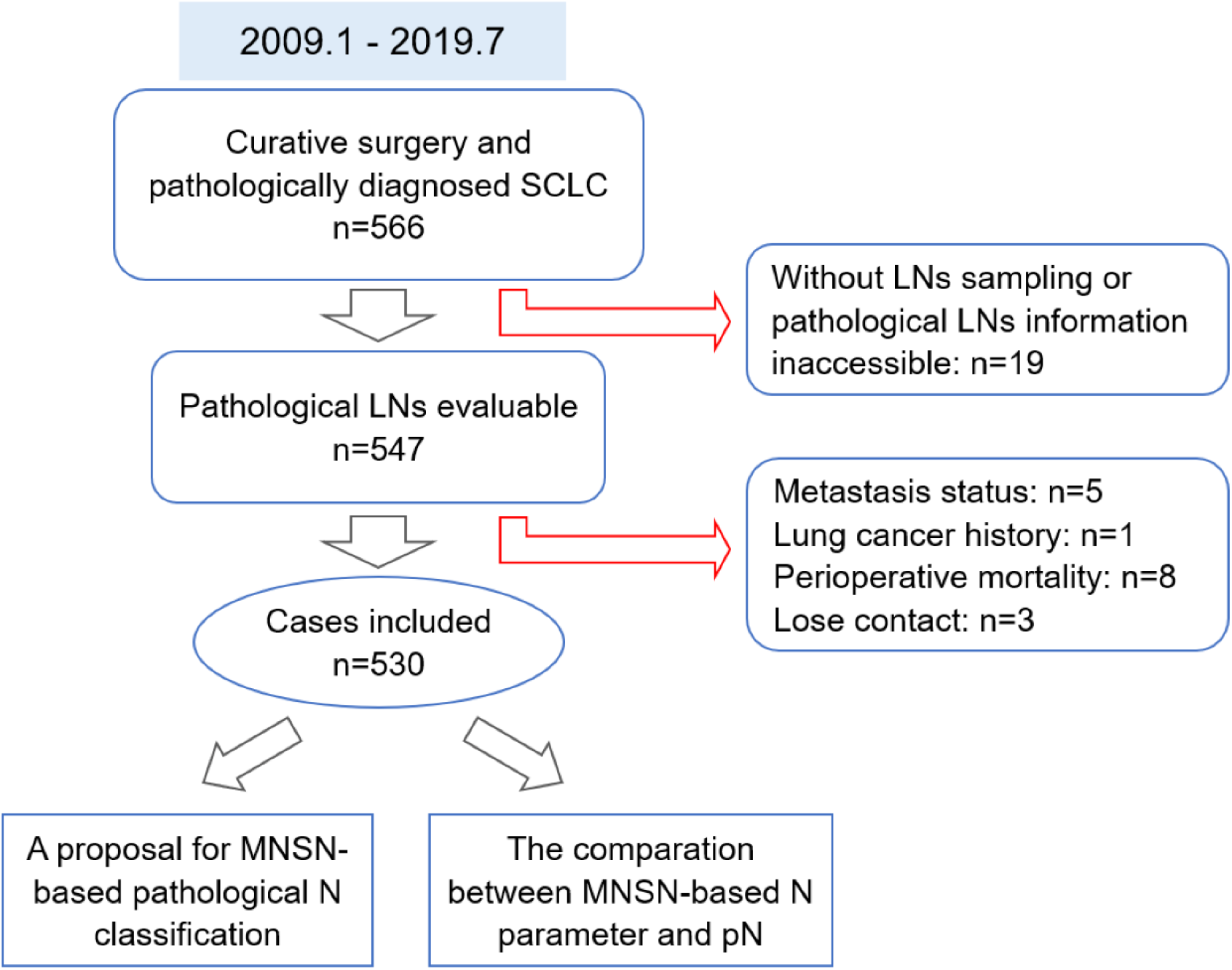
Study design. MNSN, Metastatic Lymph Node Station Number; pN, pathologic N category in TNM staging system

**Table 1.** Baseline Characteristics and Pathological Features

| Characteristic | No. | Percentage (%) |
| --- | --- | --- |
| Mean Age (y, ±SD) | 60.7 ±9.0 | |
| Sex |  |  |
| Male | 461 | 87.0 |
| Female | 69 | 13.0 |
| Smoking History |  |  |
| Yes | 278 | 52.5 |
| No | 252 | 47.5 |
| Location |  |  |
| Right Lung | 254 | 47.9 |
| Left Lung | 276 | 52.1 |
| Extent of pulmonary resection |  |  |
| Sublobar resection | 16 | 3.0 |
| Lobectomy | 353 | 66.6 |
| Bilobectomy | 70 | 13.2 |
| Pneumonectomy | 91 | 17.2 |
| Means of surgery |  |  |
| Open | 262 | 49.4 |
| VATS | 268 | 50.6 |
| Uniportal | 100 | 37.3 |
| Mean tumor size (cm, ±SD) | 3.34 ±1.77 | |
| T Status |  |  |
| Tx | 34 | 6.4 |
| T1 | 206 | 38.9 |
| T1a | 17 | 8.3 |
| T1b | 95 | 46.1 |
| T1c | 94 | 45.6 |
| T2 | 203 | 38.3 |
| T2a | 156 | 76.8 |
| T2b | 47 | 23.2 |
| T3 | 65 | 12.3 |
| T4 | 22 | 4.2 |
| Mean No. of LNs resected (±SD) | 13.2 ±6.5 | |
| Histologic subtype |  |  |
| Pure SCLC | 371 | 70.0 |
| Mixed SCLC | 159 | 30.0 |
| Visceral pleura invasion |  |  |
| Absent | 499 | 94.2 |
| Present | 31 | 5.8 |
| Nerve invasion |  |  |
| Absent | 521 | 98.3 |
| Present | 9 | 1.7 |
| Lymphatic vascular invasion |  |  |
| Absent | 482 | 90.9 |
| Present | 48 | 9.1 |
| Adjuvant treatment |  |  |
| Chemotherapy | 387 | 73.0 |
| Radiotherapy | 152 | 28.7 |

SD, standard deviation; VATS, video-assisted thoracoscopic surgery; LN, lymph node; SCLC, small cell lung cancer.

### MNSN-based LN staging predicts survival ideally

To reveal the relationship of MNSN and survival, the Kaplan-Meier method was employed, which showed that patients with greater MNSN had worse OS and RFS (Fig. 2). Meanwhile, some patients with adjacent MNSN shared similar survivals, suggesting the necessity to define groups. Hence, patients were classified into 4-5 groups according to the adjacency of MNSN and similarity of survival. Finally, two models were ideal and it could predict both OS and RFS (Fig. 3) when MNSN was classified as 0, 1–2, 3–5 and 6–7, four groups. Another grouping strategy was considered unsatisfactory as its less efficacy in predicting RFS, though it performed well in OS (see Supplementary Fig. 1).

To balance other hazard factors, sex, age, smoking, tumor purity and pathologic T stage were adjusted in multi-variate COX proportional hazards model, which suggested MNSN group was an independent hazard factor for both OS and RFS (Table 2).

**Figure 2.**
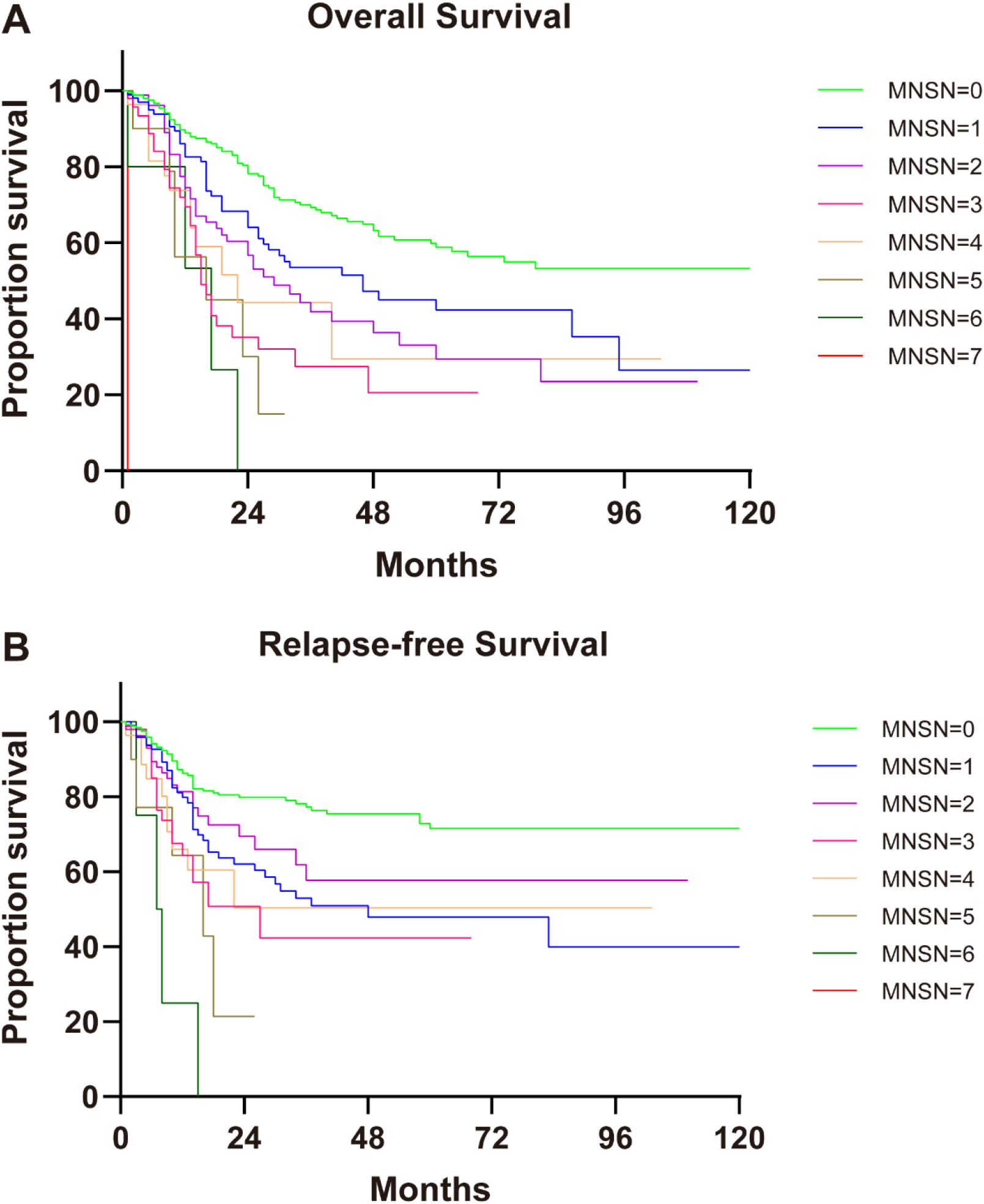
Kaplan-Meier curve for OS (**A**) and RFS (**B**) by MNSN. MNSN, Metastatic Lymph Node Station Number

**Figure 3.**
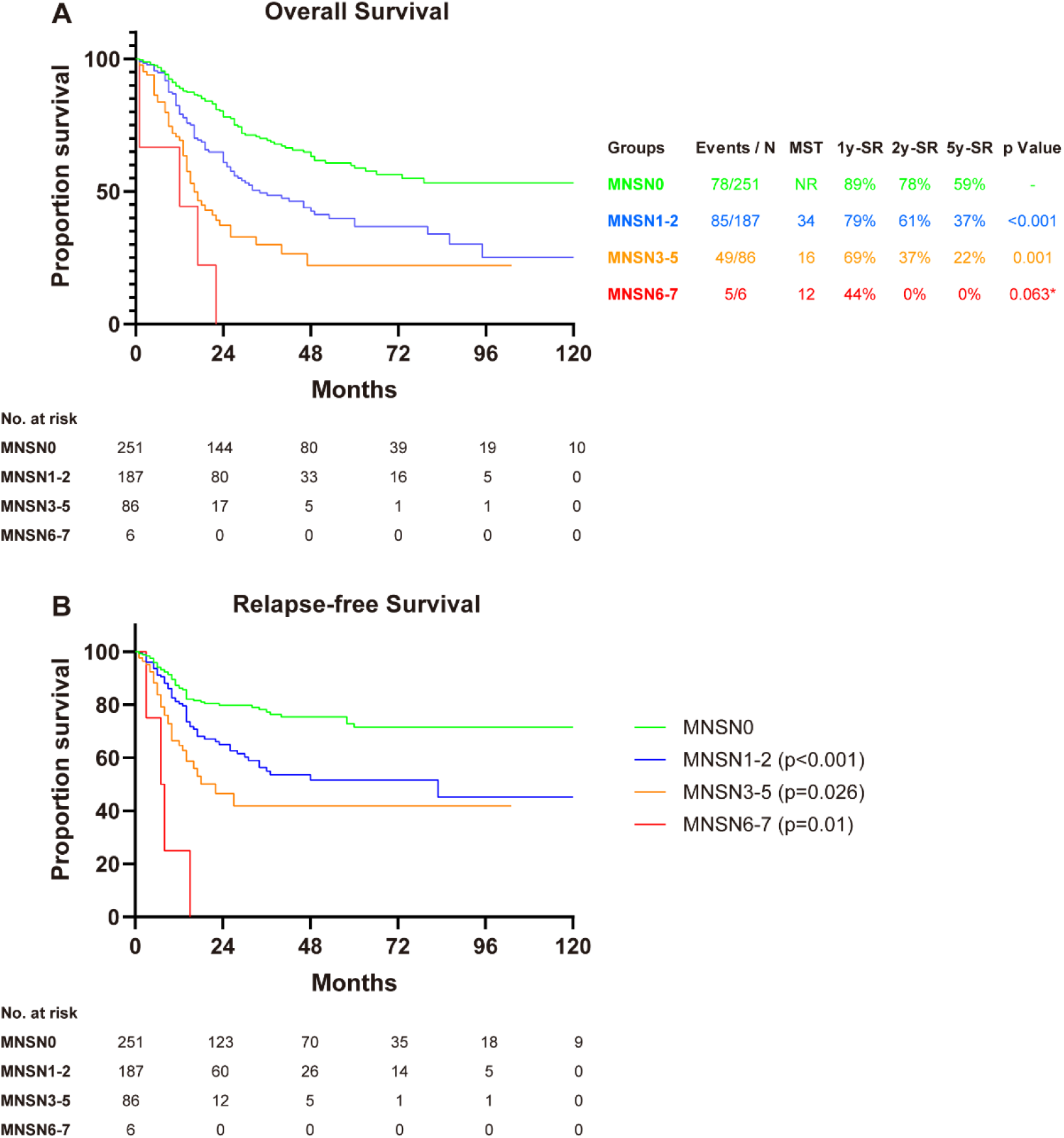
Kaplan-Meier curve for OS (**A**) and RFS (**B**) grouped by MNSN. The p value in the graph indicates the significance of log-rank test compared with the upper group.^*^The difference between MNSN6–7 and MNSN3–5 is not statistically significant, probably due to the small sample capacity in the MNSN6–7 group. OS, overall survival; RFS, recurrence-free survival; 1y-SR, 1-year survival rate, and so on; MNSN, Metastatic Lymph Node Station Number

**Table 2.** Multivariate Cox Proportional Hazards Model Analyses for the Efficacy of MNSN Groups on OS and RFS Adjusted for sex, age, smoking, tumor purity and pathological T stage

| Group | OS |  | RFS |  |
| --- | --- | --- | --- | --- |
|  | Hazard Ratio (95% CI) | p Value | Hazard Ratio (95% CI) | p Value |
| MNSN1-2<br>vs. MNSN0 | 1.923 [1.4.6-2.628] | <0.001 | 2.014 [1.382-2.934] | <0.001 |
| MNSN3-5<br>vs. MNSN0 | 3.785 [2.600-5.512] | <0.001 | 3.194 [2.039-5.004] | <0.001 |
| MNSN6-7<br>vs. MNSN0 | 7.446 [2.946-18.818] | <0.001 | 11.596 [4.143-32.457] | <0.001 |
| MNSN3-5<br>vs. MNSN1-2 | 1.969 [1.372-2.826] | <0.001 | 1.586 [1.031-2.440] | 0.036 |
| MNSN6-7<br>vs. MNSN1-2 | 3.873 [1.541-9.731] | 0.004 | 5.759 [2.077-15.963] | 0.001 |
| MNSN6-7<br>vs. MNSN3-5 | 1.967 [0.775-4.992] | 0.155* | 3.630 [1.279-10.305] | 0.015 |

MNSN: Metastatic Lymph Node Station Number; OS, overall survival; RFS, recurrence-free survival; CI, confidence interval.
\*The difference between MNSN6-7 and MNSN3-5 is not statistically significant, probably due to the small sample capacity the in the MNSN6-7 group.

### MNSN-based N parameter might be better than pN

In 2015, the analysis results of IASLC database indicated that pN in eighth edition of TNM staging system was suitable for SCLC. To compare pN with MNSN, we also analyzed its discriminatory power for OS and RFS by Kaplan-Meier curve and log-rank test (Fig. 4A-B). Although N0 stage was well distinguished from other stages for OS (N0 vs N1, p = 0.001; N0 vs N2, p< 0.001), N1 stage overlapped with N2 stage (p = 0.099). As for RFS, N2 stage also couldn’t be separated from N1 stage (p = 0.254). The discriminatory power of pN for N1 and N2 stage in our cohort was similar with it in SEER database (Fig. 4C), in which the survival curves totally overlapped (p = 0.413).

**Figure 4.**
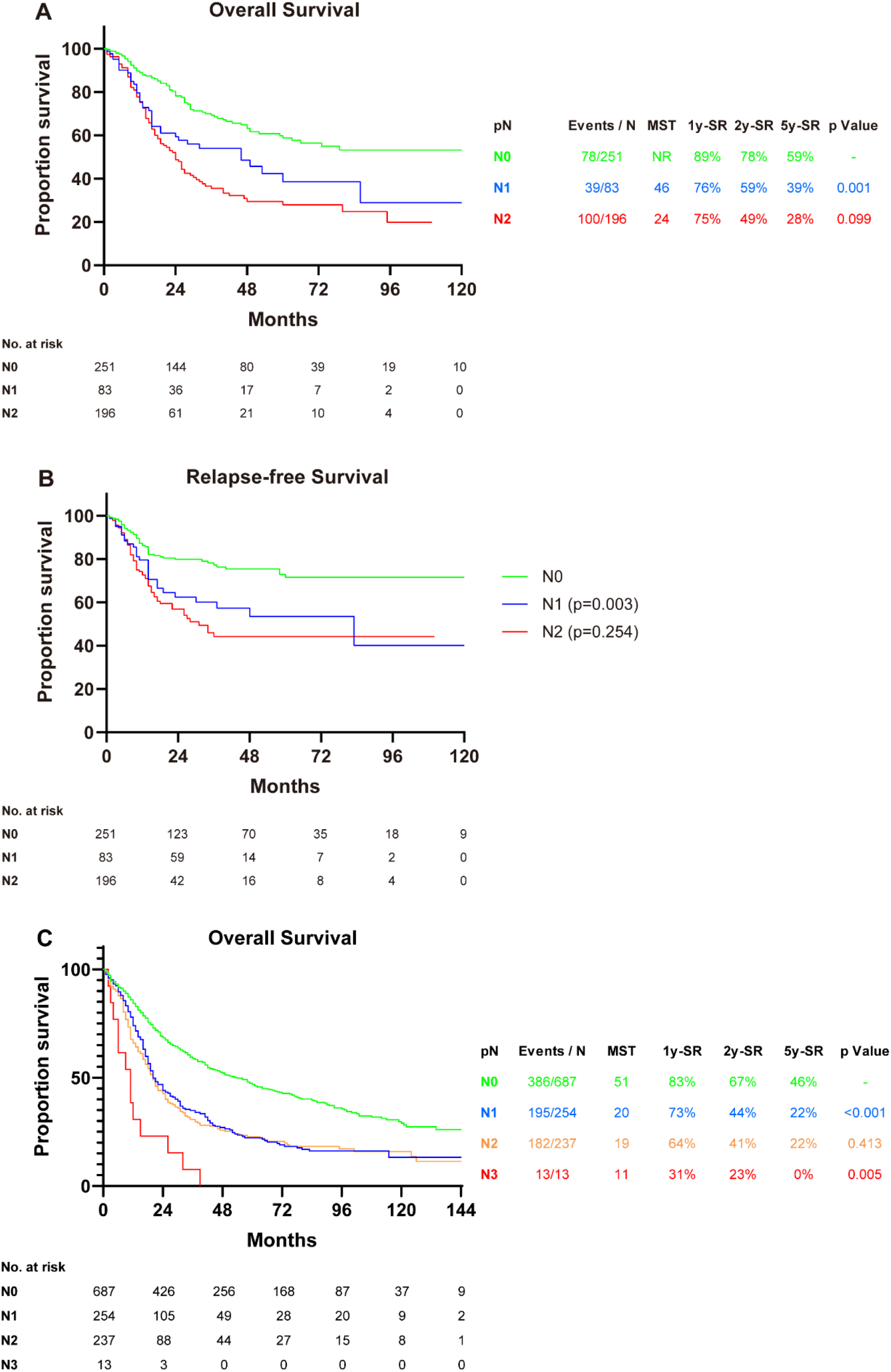
Kaplan-Meier curve for OS and RFS by the N category in eighth edition of TNM staging system using data from our cohort (**A**-**B**) or from the Surveillance, Epidemiology, and End Results database (**C**). The p value in the graph indicates the significance of log-rank test compared with the upper group. OS, overall survival; RFS, recurrence-free survival; 1y-SR, 1-year survival rate, and so on.

## Discussion

The currently used pathologic N parameter for lung cancer in 8th edition of TNM staging system was identical to 7th edition, which was proposed in 2009 and had not been changed thereafter^8^. Though it was validated to be effective in predicting survival, controversies existed as for its discriminatory ability of the N1 and N2 stage.

In NSCLC, several studies revealed the heterogeneity in N1 and N2 category^23,24^. Therefore, subdividing of N1 and N2 into more groups was put forward. It was suggested that N1 could be divided into N1a, N1b and N2 could be divided into N2a1, N2a2, N2b according to the number of involved lymph node stations and the presence of skip metastasis. However, analyses of the most updated IASLC lung cancer database indicated the overlap of survival curve in N1b and N2a2, and N2a1 tended to be better than N1b, though not significant. Thus, the IASLC suggested N parameter should be maintained in the 8th staging system^11^. Other studies recommending the combination of pN with the number of involved lymph node stations^20,25^ or induction of lymph node ration (LNR)^22^ were worth considering, but should be further validated.

In contrast, study of regrouping strategy for pathologic N parameter in SCLC was rare. The IASLC had launched 2 studies evaluating the efficacy of pathologic N parameter in 7th or 8th TNM system in SCLC. The first was carried out in 2009, containing 349 surgical cases in 1990-2000 where pathologic TNM staging was available. In that database, survival curves were separated from each other, though N0 overlapped with N1 in the tail, and the median survival time was 51, 24, 13 and 6 months for N0 to N3 groups^8^. Differently, the database in 2015 containing 582 patients with adequate pathologic stages in 1999-2009 showed a significant difference between N0 and N1 whereas less difference between N1 and N2^10^. The disagreement of the 2 studies was worth exploring. Since the cases from the later study had no overlap with the former one and were a decade lagging behand, the surgical, pathological and therapeutic improvements may account for the discordance, awaiting more up to date data to verify.

In this study, we established a database containing 566 patients who were operated and pathologically confirmed SCLC in our hospital during 2009-2019. Among those patients, 530 were eligible for the evaluation of N parameter. Consistent with the most updated IASLC database and SEER database, our data showed remarkable difference between N0 and N1 but little even no statistical difference between N1 and N2. By contrast, the metastatic lymph node station number proposed by us was closely related to prognosis. Besides, when MNSN was classified into several groups, it could predict survival better. The result that number of involved lymph node station had preferable predicting value may be explained. The SCLC is a histological subtype characterized by rapid local and distal metastasis. Hence, the location and the distance of regional lymph nodes in SCLC may not be as important as those in NSCLC. Instead, the number of involved lymph node stations which represent the invasiveness and the number of tumor cells to further disseminate distally may play a major role.

Some studies nominally referring to the number of involved lymph node stations were actually the combination of location of metastatic nodes, single station versus multiple stations and absence versus presence of skip metastasis^15,20,21^. In other words, to our knowledge, this is the first study which merely use the number of involved lymph node stations independent of node location to predict survival in lung cancer. Accordingly, biases exist in our study including the retrospective nature, the loss to follow-up rate, the rationality of grouping strategy, et al. The median follow-up time is 21 months in our cohort, which seems to be inadequate. However, considering that the median survival time in SCLC was about 15 months^26^, it is sufficient to observe expected outcomes. Nevertheless, more sophisticatedly designed studies are acclaimed to verify this parameter.

In a word, we proposed a novel pathologic N parameter based on metastatic lymph node station number and it might be a better indicator to predict survival than currently used pN in SCLC. The reliability of this parameter should be further validated in large-scale, prospective studies.

## Data Availability

The raw/processed data required to reproduce these findings cannot be shared at this time as the data also forms part of an ongoing study.

## Acknowledgments

Dr. Peng Zhang, Dr Gening Jiang, and Dr. Yuming Zhu contributed equally to the study. This study was supported by the National Natural Science Foundation of China [Grant No. 81972172], the Shanghai Hospital Development Center [Grant No. SHDC12018122], the Shanghai Science and Technology Committee [Grant No. 19XD1423200], and Programs of Shanghai Pulmonary Hospital [No. fkcx1904].

## References

1. Bray F, Ferlay J, Soerjomataram I, et al: Global cancer statistics 2018: GLOBOCAN estimates of incidence and mortality worldwide for 36 cancers in 185 countries. CA: a cancer journal for clinicians 68: 394–424, 2018

2. Navada S, Lai P, Schwartz AG, et al: Temporal trends in small cell lung cancer: Analysis of the national Surveillance, Epidemiology, and End-Results (SEER) database. Journal of clinical oncology: official journal of the American Society of Clinical Oncology 24: 7082–7082, 2006

3. Govindan R, Page N, Morgensztern D, et al: Changing epidemiology of small-cell lung cancer in the United States over the last 30 years: analysis of the surveillance, epidemiologic, and end results database. Journal of clinical oncology: official journal of the American Society of Clinical Oncology 24: 4539–4544, 2006

4. Shields TW, Higgins GA, Matthews MJ, et al: Surgical resection in the management of small cell carcinoma of the lung. The Journal of thoracic and cardiovascular surgery 84: 481–488, 1982

5. Zelen M: Keynote address on biostatistics and data retrieval. Cancer chemotherapy reports. Part 3 4: 31–42, 1973

6. Stahel RA, Ginsberg R, Havemann K, et al: Staging and prognostic factors in small cell lung cancer: a consensus report. lung cancer 5: 119–126, 1989

7. Shepherd FA, Crowley J, Van Houtte P, et al: The International Association for the Study of Lung Cancer lung cancer staging project: proposals regarding the clinical staging of small cell lung cancer in the forthcoming (seventh) edition of the tumor, node, metastasis classification for lung cancer. Journal of thoracic oncology: official publication of the International Association for the Study of lung cancer 2: 1067–1077, 2007

8. Vallières E, Shepherd FA, Crowley J, et al: The IASLC Lung Cancer Staging Project: proposals regarding the relevance of TNM in the pathologic staging of small cell lung cancer in the forthcoming (seventh) edition of the TNM classification for lung cancer. Journal of thoracic oncology: official publication of the International Association for the Study of lung cancer 4: 1049–1059, 2009

9. Rami-Porta R, Asamura H, Travis WD, et al: Lung cancer - major changes in the American Joint Committee on Cancer eighth edition cancer staging manual. CA: a cancer journal for clinicians 67: 138–155, 2017

10. Nicholson AG, Chansky K, Crowley J, et al: The International Association for the Study of Lung Cancer Lung Cancer Staging Project: Proposals for the Revision of the Clinical and Pathologic Staging of Small Cell Lung Cancer in the Forthcoming Eighth Edition of the TNM Classification for Lung Cancer. Journal of thoracic oncology: official publication of the International Association for the Study of Lung Cancer 11: 300–311, 2016

11. Asamura H, Chansky K, Crowley J, et al: The International Association for the Study of Lung Cancer Lung Cancer Staging Project: Proposals for the Revision of the N Descriptors in the Forthcoming 8th Edition of the TNM Classification for Lung Cancer. Journal of thoracic oncology: official publication of the International Association for the Study of Lung Cancer 10:1675–1684, 2015

12. Andre F, Grunenwald D, Pignon JP, et al: Survival of patients with resected N2 non-small-cell lung cancer: evidence for a subclassification and implications. Journal of clinical oncology: official journal of the American Society of Clinical Oncology 18: 2981–2989, 2000

13. Asamura H, Suzuki K, Kondo H, et al: Where is the boundary between N1 and N2 stations in lung cancer? The Annals of thoracic surgery 70, 2000

14. Jonnalagadda S, Smith C, Mhango G, et al: The number of lymph node metastases as a prognostic factor in patients with N1 non-small cell lung cancer. Chest 140: 433–440, 2011

15. Lee JG, Lee CY, Park IK, et al: Number of metastatic lymph nodes in resected non-small cell lung cancer predicts patient survival. The Annals of thoracic surgery 85: 211–215, 2008

16. Wisnivesky JP, Arciniega J, Mhango G, et al: Lymph node ratio as a prognostic factor in elderly patients with pathological N1 non-small cell lung cancer. Thorax 66: 287–293, 2011

17. Qiu C, Dong W, Su B, et al: The prognostic value of ratio-based lymph node staging in resected non-small-cell lung cancer. Journal of thoracic oncology: official publication of the International Association for the Study of Lung Cancer 8: 429–435, 2013

18. Lee JG, Lee CY, Bae MK, et al: Validity of International Association for the Study Of Lung Cancer proposals for the revision of N descriptors in lung cancer. Journal of thoracic oncology: official publication of the International Association for the Study of Lung Cancer 3: 1421–1426, 2008

19. Rusch VW, Crowley J, Giroux DJ, et al: The IASLC Lung Cancer Staging Project: proposals for the revision of the N descriptors in the forthcoming seventh edition of the TNM classification for lung cancer. Journal of thoracic oncology: official publication of the International Association for the Study of Lung Cancer 2: 603–612, 2007

20. Park BJ, Kim TH, Shin S, et al: Recommended Change in the N Descriptor Proposed by the International Association for the Study of Lung Cancer: A Validation Study. Journal of thoracic oncology: official publication of the International Association for the Study of lung cancer 14: 1962–1969, 2019

21. Saji H, Tsuboi M, Shimada Y, et al: A proposal for combination of total number and anatomical location of involved lymph nodes for nodal classification in non-small cell lung cancer. Chest 143: 1618–1625, 2013

22. Ding X, Hui Z, Dai H, et al: A Proposal for Combination of Lymph Node Ratio and Anatomic Location of Involved Lymph Nodes for Nodal Classification in Non-Small Cell Lung Cancer. Journal of thoracic oncology: official publication of the International Association for the Study of lung cancer 11: 1565–1573, 2016

23. Kang CH, Ra YJ, Kim YT, et al: The impact of multiple metastatic nodal stations on survival in patients with resectable N1 and N2 nonsmall-cell lung cancer. The Annals of thoracic surgery 86: 1092–1097, 2008

24. Shimada Y, Tsuboi M, Saji H, et al: The prognostic impact of main bronchial lymph node involvement in non-small cell lung carcinoma: suggestions for a modification of the staging system. The Annals of thoracic surgery 88: 1583–1588, 2009

25. Chen W, Zhang C, Wang G, et al: Feasibility of nodal classification for non-small cell lung cancer by merging current N categories with the number of involved lymph node stations. Thoracic cancer 10: 1533–1543, 2019

26. Schabath MB, Nguyen A, Wilson P, et al: Temporal trends from 1986 to 2008 in overall survival of small cell lung cancer patients. Lung Cancer 86: 14–21, 2014

